# Understanding the Feasibility of Computer Vision in Diagnosing Respiratory Infections in Pediatric Emergency Rooms

**DOI:** 10.1101/2025.04.17.25325898

**Authors:** Ella Jokinen, Ella Pääkkönen, Constantino Álvarez Casado, Aino K. Rantala, Terhi Ruuska-Loewald, Jouni J.K. Jaakkola, Miguel Bordallo López

**Affiliations:** Center for Environmental and Respiratory Health Research, Research Unit of Population Health, University of Oulu; Center for Machine Vision and Signal Analysis, University of Oulu; Department of Pediatrics and Adolescent Medicine, Medical Research Center, Oulu University Hospital, and Research Unit of Clinical Medicine, University of Oulu; Finnish Meteorological Institute, Helsinki, Finland; Biocenter Oulu, University of Oulu, Finland

**Keywords:** pediatric emergency care, respiratory infections, respiratory condition assessment, computer vision diagnostics, non-intrusive monitoring, machine learning in healthcare

## Abstract

**Objective:** Respiratory infections are a leading cause of pediatric emergency visits globally, requiring timely and accurate assessment. This study evaluated the feasibility of a computer vision-based system to identify respiratory infections or distress and estimate vital signs, including respiratory rate (RR), heart rate (HR), and oxygen saturation (SpO2), in pediatric patients visiting an emergency department.

**Methods:** We conducted a population-based case-control study involving 100 children aged 0–5 years at the Children’s Emergency Department of Oulu University Hospital, Finland, between April and June 2022. Cases included children with respiratory symptoms (n=64), while controls had no respiratory symptoms (n=36). High-quality videos were recorded using a smartphone while the children were sitting on the lap of a parent, capturing the facial and precordium regions. Biosignals were extracted from the videos using remote photoplethysmography (rPPG) and remote ballistography (rBSG). From these 1D signals we calculated 250 features per window, including statistical, respiratory and heart-related features. These features were used to train machine learning models for classifying respiratory conditions and estimating vital signs. Ground truth data included manually measured SpO2, HR, RR, and body temperature, while nasopharyngeal samples were analyzed to determine infection etiology.

**Results:** The model achieved a classification accuracy of 86% in predicting SpO2 levels above 95 and 73% for levels below 95. It classified dyspnea with 63% of balanced accuracy and respiratory infections with 77% of balanced accuracy. For predicting hospitalization, the classification accuracy was 67%. Challenges were noted in video analysis due to patient movement and age-related variability, with better signal quality observed in sicker patients who remained still.

**Conclusions:** This study demonstrates the potential of computer vision systems to provide automated, non-intrusive assessments of respiratory conditions in acutely ill children. This could enable even remote assessment of acutely ill children at home. While the results show moderate accuracy, further research is needed to improve reliability across diverse clinical scenarios and patient populations.

## 1. Introduction

Respiratory infections are a leading cause of mortality, morbidity, and emergency department visits in children [1]–[3]. Early diagnosis and timely management of respiratory infections are critical to improving patient outcomes, especially in young children, who are more vulnerable to serious complications. In clinical practice, assessments often rely on vital signs, such as respiratory rate (RR), heart rate (HR), and oxygen saturation (SpO2), as well as scoring systems such as the Respiratory Stress Assessment Index (RDAI) and the Pediatric Early Warning Score (PEWS) [4], [5]. Although these tools are indispensable, traditional methods often involve manual measurements and subjective evaluations, which can be inconsistent and time-consuming, particularly in high-pressure emergency settings [6], [7]. Furthermore, these methods are not suitable for remote assessment of children at home because the parental ability to assess pediatric vital signs is limited [8].

Advances in digital health technologies, particularly computer vision and artificial intelligence, offer promising alternatives for augmenting diagnostic capabilities in health care, including remote assessment of acutely ill children through digital services [9]. Non-contact diagnostic systems, which analyze visual or radio frequency data to extract physiological signals, have demonstrated potential in areas such as sleep monitoring, recognition of stress and depression, and neonatal care [10], [11]. Computer vision methods provide a non-intrusive means of assessing vital signs by leveraging techniques such as remote photoplethysmography (rPPG) and remote ballistocardiography (rBSG) to measure subtle variations in skin color and motion, respectively [12], [13]. These systems could address the limitations of traditional methods by providing automated, standardized, and efficient evaluations of respiratory distress and related conditions.

Many studies have explored non-contact vital sign monitoring using these novel technologies, such as rPPG with RGB cameras, rBCG with radars, and respiratory monitoring through RGB, depth cameras, and radar systems, including camera-guided radars. These approaches have demonstrated reliable detection of heart rate, respiratory rate, and sleep patterns, particularly in controlled settings such as sleep laboratories, intensive care units, or other static environments [10], [12]– [16]. Multimodal techniques that integrate motion tracking and rPPG signals have achieved clinically acceptable accuracy in respiratory monitoring tasks, such as a mean absolute error of 1.33 breaths per minute for respiratory rate estimation [17]. Despite these advances, most of the datasets and studies involve adult populations, static users, and highly constrained conditions, which minimize external variability. As a result, these methods often fail to address the challenges posed by dynamic environments in the real world such as pediatric emergency departments, where movement, unpredictable behavior, and physiological variability are significant [18]–[20]. Expanding these technologies to pediatric settings requires further research to ensure their applicability and reliability in such complex scenarios [10], [20].

Our study addressed this gap by evaluating a computer vision-based system to monitor respiratory and cardiovascular parameters in children presenting to a pediatric emergency department. Unlike previous research confined to static or controlled environments, this study emphasizes the practical challenges and variability inherent in real-world clinical settings. By applying camera-based technologies to pediatric patients with respiratory infections or distress, our objective was to assess the feasibility, accuracy, and limitations of this approach. In addition, our study aims to bridge the gap between experimental methods and their clinical applicability and to offer insight into the potential of non-contact monitoring in dynamic healthcare environments. The key elements of the study were:

- Evaluation in a Real-World Pediatric Emergency Department: We conducted this study in a dynamic clinical setting, addressing practical challenges such as patient movement and variability under ambient conditions.
- Comprehensive Signal Analysis: The proposed system integrates rPPG and rBSG techniques to extract both color and motion signals from video recordings, enabling the estimation of multiple vital signs, including RR, HR, and SpO2.
- Focus on Pediatric Populations: The study emphasizes the unique challenges and requirements of assessing young children, providing insights into the applicability of non-contact methods in pediatric care.
- Integration of Clinical Metrics: In addition to estimating vital signs, we utilized scoring systems such as RDAI and PEWS to evaluate respiratory distress and the need for hospitalization, enhancing the clinical relevance of our findings.
- Identification of Practical Challenges: By analyzing performance across diverse patient groups and conditions, we highlight key limitations and areas for improvement in the implementation of video-based diagnostic systems.

This work fundamentally advances the integration of non-contact monitoring technologies in pediatric emergency care by addressing both clinical and technical challenges in real-world settings. Its findings provide a foundation, especially for identifying challenges and opportunities, while opening avenues for further research to enhance the applicability and reliability of these technologies.

The primary objective of this study was to evaluate whether computer vision can accurately identify patients with a respiratory infection or respiratory distress in a pediatric emergency department. Furthermore, we assessed how accurately computer vision estimates vital signs such as RR, HR, SpO2, dyspnea, and the need for hospitalization in acutely ill children.

## 2. Methodology

### 2.1. Study design and population

This was a population-based incident case-control study of respiratory infections in children entering the Emergency Department of Oulu University Hospital in Oulu, Northern Finland, serving a geographically defined region between April and June 2022. The Regional Medical Research Ethics Committee of the Wellbeing Services County of North Ostrobothnia approved the study protocol.

The Emergency Department treats all acute cases from a geographically defined region. We invited a total of 145 children aged 0-5 years and their parents upon arrival in the pediatric ED (see Figure 1 for the selection of the study population). Of these, 107 agreed to participate (73.8%). All parents or guardians gave their written informed consent. We obtained 82 participants with two videos, 18 patients with one video (11 sitting and 7 lying down), and 7 patients were excluded from the study due to low video quality. Non-Finnish speakers were excluded from the study. The final study population consisted of 100 children, 64 cases whose main reason for seeking treatment was respiratory infections, and 36 controls with no respiratory symptoms. Both groups were divided into two age groups: 0-2 years old and 2-5 years old.

**Figure 1.**
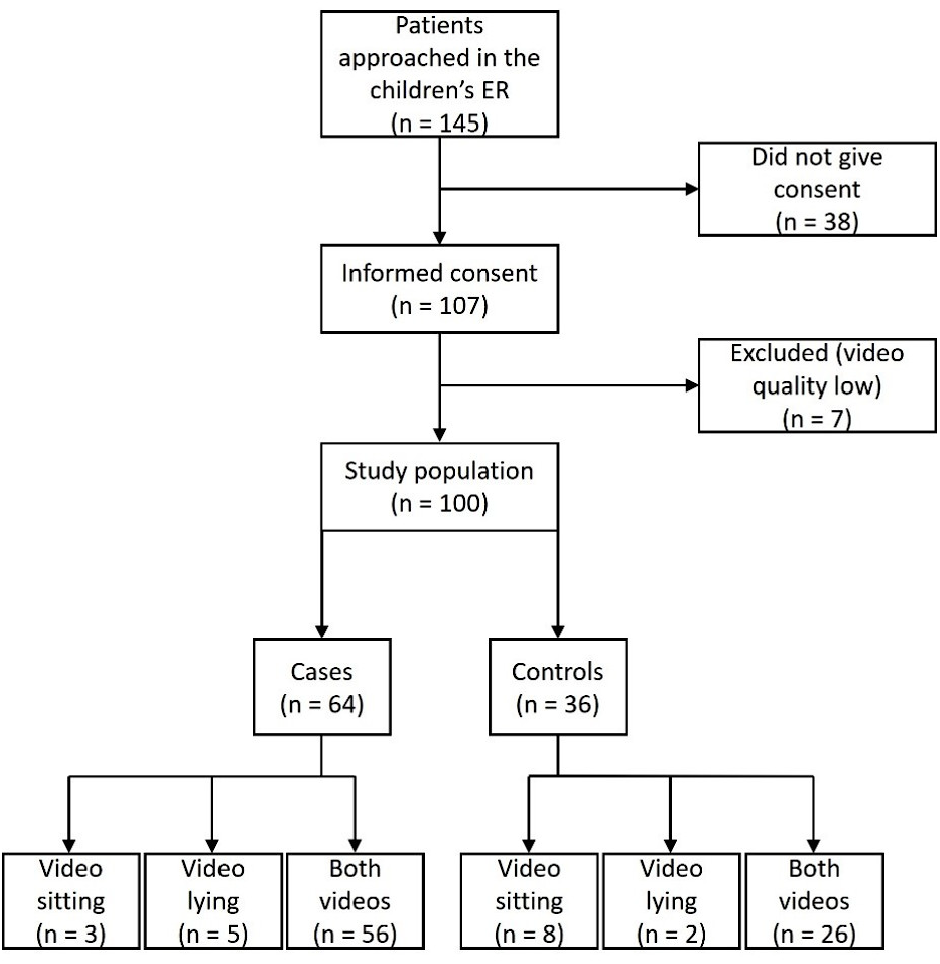
Study design.

### 2.2. The clinical evaluation of the study participants

The study physicians collected information on the child’s clinical characteristics from medical records. In addition, the guardians filled out a questionnaire about the child’s previous respiratory infections, asthma and allergies, pets at home, and antenatal and postnatal exposure to tobacco smoke.

An emergency department (ED) nurse measured the child’s weight and height, SpO2 and HR (measured with a fingertip pulse oximeter), and body temperature using a digital thermometer. RR was calculated manually by examining breathing movements either by an ED nurse, an ED physician or the research assistants.

An ED physician evaluated dyspnea and other clinical symptoms measured by the RDAI and PEWS. Dyspnea, or shortness of breath, characterized by wheezing, lowered oxygen saturation and use of accessory muscles in respiration, was used to describe the symptoms of the patients. The patient had dyspnea if one or multiple measurements evaluating breathing were significantly lower than average. RDAI score is formed from the physician’s analysis of wheezing and retractions, and it estimates deviances in respiratory status [21]. PEWS score is an age-related index that targets to identify patients at risk of clinical deterioration, and is formed by RR, breathing difficulty, SpO2, need for supplemental oxygen, systolic blood pressure, HR, capillary function, and consciousness level [22]. In this study, we estimated the PEWS score with only the available measurements (RR, breathing difficulty, SpO2, HR and consciousness). In addition, the viral and bacterial etiology of respiratory infections was analyzed from nasopharyngeal samples by polymerase chain reaction (PCR) or antibody testing. Nasophrayngeal swabs for multiplex PCR to detect respiratory pathogens were obtained as part of clinical routine practice.

### 2.3 Video recording and data capturing

Two 30-second videos of each patient were recorded by study physicians in the pediatric ED. One video in which the child sits down, usually on the parent’s lap, was recorded with the smartphone placed on a tripod. Another video of the child lying down was recorded with the smartphone held by hand. The videos were recorded using similar lighting in the same room with a similar distance from the patient.

### 2.4. Computer vision methodology for Pediatric Medical Diagnosis Support

We conducted an automatic analysis of the recorded videos to support the diagnosis of respiratory infections. We leveraged state-of-the-art computer vision and signal processing techniques to extract respiratory and blood pulse biosignals from the face and torso of patients and used machine learning to classify respiratory conditions using computed respiratory and heart-related features extracted from the biosignals.

The video analysis methodology consisted of five main stages, as depicted in Figure 2. First, we captured high-quality video streams of patients using a Nokia X10 smartphone with a rear camera at a resolution of 1920x1080 and 30 frames per second (FPS). Second, we performed detection, tracking, and skin segmentation to isolate the relevant regions of the face and precordium. Third, we extracted a total of four biosignals from these regions by analyzing subtle variations in color, i.e. remote photoplethysmography (rPPG) or motion, i.e. remote ballistography (rBSG). Then, we derived respiratory and heart-related features from the extracted biosignals. Subsequently, these features were used to train machine learning models for both classification and regression tasks related to respiratory conditions. Specifically, we applied models including XGBoost, Random Forest (RF), Support Vector Machines (SVM), and Multi-Layer Perceptron (MLP), following an approach similar to [23].

**Figure 2.**
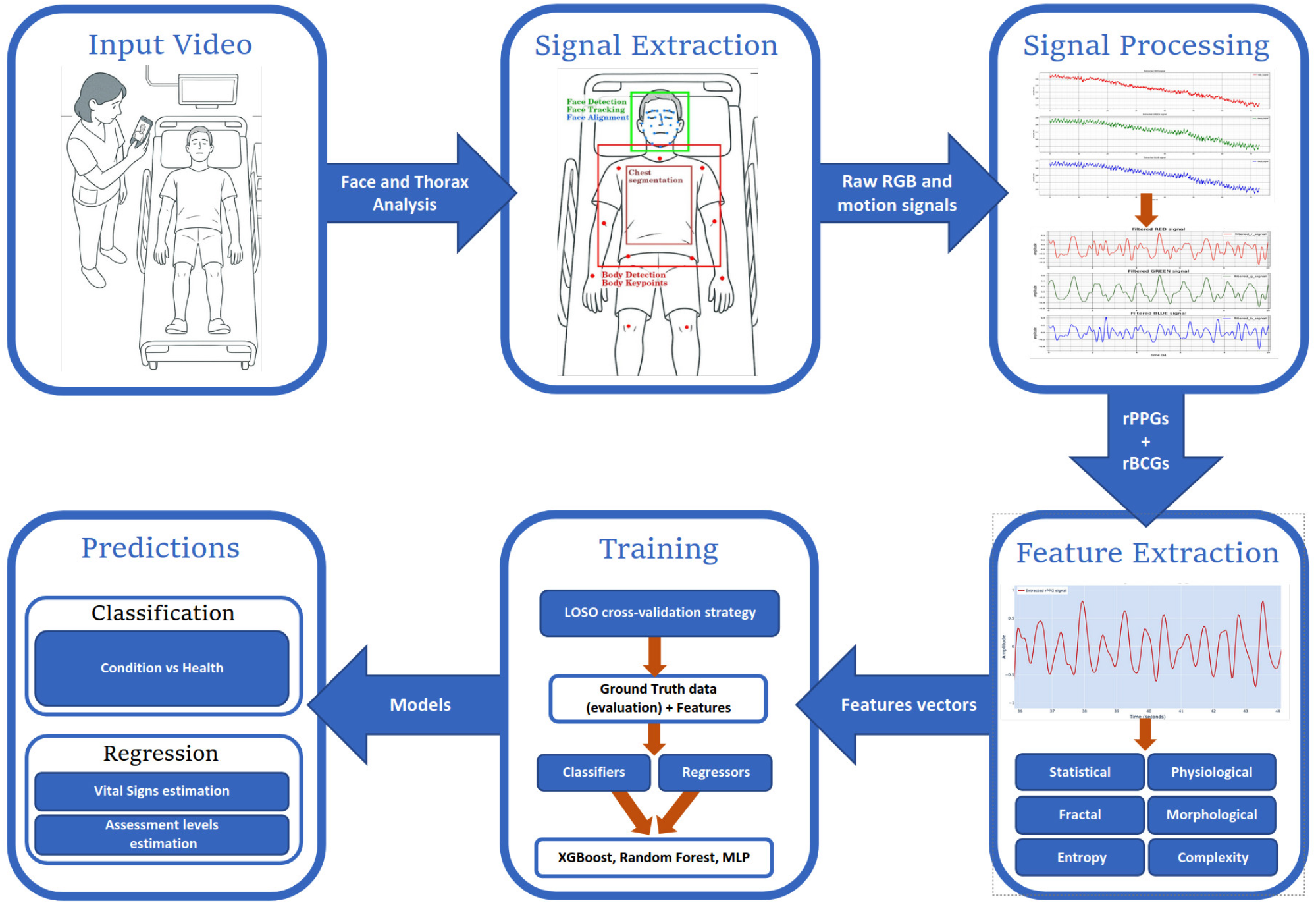
Schematic representation of the computer vision-based pipeline for video analysis and prediction of respiratory conditions. The process begins with the capture of input video, from which facial and precordial regions are analyzed to extract physiological signals using remote photoplethysmography (rPPG) and remote ballistography (rBSG). These signals are processed and used to compute a set of features, including statistical, physiological, morphological, entropy, and complexity-based descriptors. The resulting feature vectors are then used to train machine learning models for classification and regression tasks, such as predicting respiratory conditions and estimating vital signs.

From both the facial and precordium areas, color signals were extracted by averaging the RGB channels at each frame throughout the video [24], while motion signals were obtained by computing the intensity and direction of the movements of facial and body key points. This resulted in four separate signals. An example of the signals can be seen in Figure 3. Each signal was divided into 18 to 25 overlapping windows, and we calculated statistical, physiological (such as heart rate variability (HRV) and breathing patterns), morphological, complexity, entropy, and fractal features.

**Figure 3.**
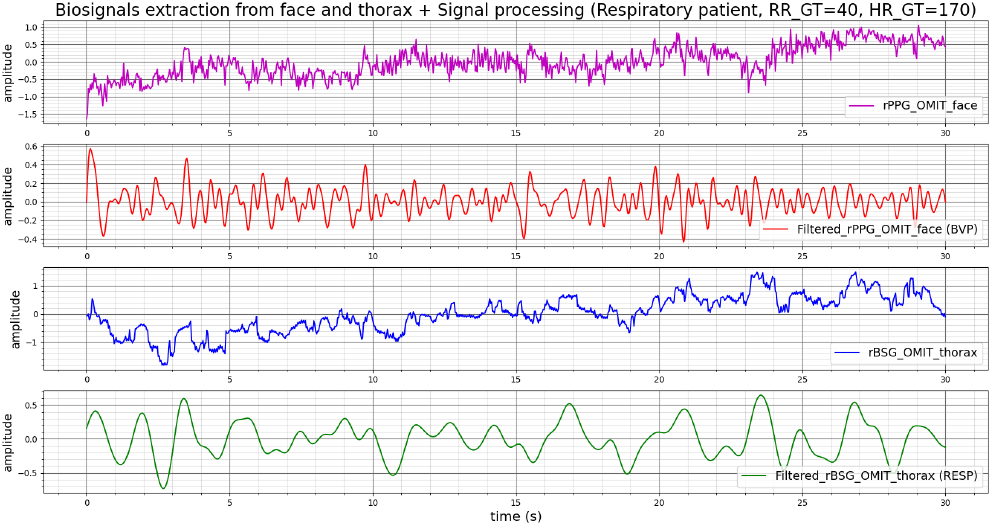
The figure displays example biosignals extracted from the face and thorax regions. The top signal shows the remote photoplethysmography (rPPG) unfiltered signal from the face, while the second signal represents the filtered blood volume pulse (BVP) signal. The third signal illustrates the remote ballistocardiography (rBSG) unfiltered signal from the thorax, and the bottom signal displays the filtered breathing signal.

### 2.5. Evaluation metrics and protocol

To obtain the predicted results, the extracted features were utilized to train a set of classifiers or regressors, using a Leave-One-Subject-Out (LOSO) validation scheme. We then assessed the performance of these models across various tasks.

To classify patients into different groups, we evaluated the performance of our computer vision methods by calculating the mean accuracy and the balanced accuracy. Mean accuracy gives a general measure of correct predictions, but in datasets with class imbalances, it may be misleading. Balanced accuracy provides a more nuanced assessment by considering the true positive rate for each class and averaging them [25]. We also visualized the results using confusion matrices. Confusion matrices offer a detailed breakdown of predictions, illustrating true positives, true negatives, false positives, and false negatives.

To evaluate the performance of our regression models, we adopted the following statistical metrics that we express in terms of Respirations per Minute (RPM), Beats per Minute (BPM), and Oxygen Saturation Percentage when estimating RR, HR, and SpO2, respectively:

- **Mean Absolute Error (MAE)**: Average absolute error between predictions and actual values, expressed as:

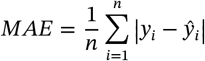

where 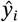 are the predicted values from our model, and y_i_ are the actual measurements taken during the doctor’s examination. Each MAE value was also documented with its standard deviation to underscore variability across subjects.
- **Mean Absolute Percentage Error (MAPE)**: Average relative error in percentage form, expressed as:

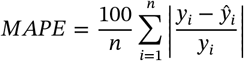
- **Root Mean Square Error (RMSE)**: Magnitude of the model’s prediction error, expressed as:

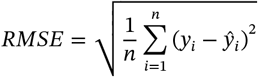
- **Coefficient of Determination (***R*^2^): Proportion of variance in the dependent variable explained by the independent variables, expressed as:

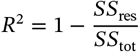

where *SS*_res_ represents the sum of the squares of the residuals (the differences between the observed and predicted values), expressed as:

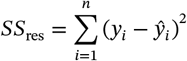

and *SS*_tot_ represents the total variance observed in the data, expressed as:

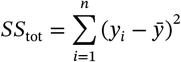

Where 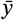 is the mean of the actual values.

We show the results in Table 2, where our findings show notable accuracy in video-based vital sign estimations, suggesting its potential applicability in specific clinical settings [26].

## 3. Results

### 3.1. Study population

Table 1 summarizes the demographic and clinical characteristics of the study population, which consisted of 64 cases (children with respiratory infections) and 36 controls (children without respiratory infections), totaling 100 children.

**Table 1.** Characteristics of the study population

|  | **Cases (n=64, 64%)** | **Controls (n=36, 36%)** | **All (n=100, 100%)** |
| --- | --- | --- | --- |
| **Age, mean (SD)** | 2.2 (1.77) | 2.6 (1.71) | 2.4 (1.75) |
| **Sex, n (%)** |  |  |  |
| Girl | 27 (42.2%) | 24 (66.7%) | 51 (51.0%) |
| Boy | 37 (57.8%) | 12 (33.3%) | 49 (49.0%) |
| **Asthma, n (%)** |  |  |  |
| No | 60 (93.8%) | 34 (94.4%) | 94 (94.0%) |
| Symptoms but not confirmed with spirometry | 3 (4.7%) | 2 (5.6%) | 5 (5.0%) |
| Yes | 1 (1.6%) | 0 (0%) | 1 (1.0%) |
| **Maternal smoking, n (%)** |  |  |  |
| No | 59 (92.2%) | 33 (91.7%) | 92 (92.0%) |
| Yes | 4 (6.3%) | 2 (5.7%) | 6 (6.0%) |
| Missing | 1 (1.6%) | 1 (2.8%) | 2 (2.0%) |
| **Reason for seeking treatment/main symptom, n (%)** |  |  |  |
| Respiratory infection | 64 (100%) | 0 (0.0%) | 64 (64.0%) |
| Stomach symptoms | 0 (0%) | 12 (33.3%) | 12 (12.0%) |
| Fever without respiratory symptoms | 0 (0%) | 2 (5.6%) | 2 (2.0%) |
| Trauma | 0 (0%) | 3 (8.3%) | 3 (3.0%) |
| Other infection | 0 (0%) | 12 (33.3%) | 12 (12.0%) |
| Control visit or measurement | 0 (0%) | 4 (11.1%) | 4 (4.0%) |
| Other | 0 (0%) | 3 (8.3%) | 3 (3.0%) |
| **RDAI score, mean (SD)** | 2.47 (3.16) | 0.06 (0.33) | 1.60 (2.79) |
| **PEWS score, mean (SD)** | 1.72 (1.97) | 0.29 (0.75) | 1.21 (1.79) |
| **Oxygen saturation, mean (SD)** | 96.9 (2.26) | 98.8 (1.32) | 97.4 (2.22) |
| **Respiratory rate, mean (SD)** | 42.3 (11.27) | 32.2 (9.67) | 38.9 (11.73) |
| **Heart rate, mean (SD)** | 139.3 (24.44) | 123.3 (32.19) | 136.1 (26.65) |
| **Body temperature, mean (SD)** | 37.8 (1.02) | 37.3 (0.68) | 37.7 (0.97) |
| **Respiratory symptoms from medical records, n (%)** |  |  |  |
| Cough | 36 (56.2%) | 1 (2.8%) | 37 (37.0%) |
| Stuffy nose | 22 (34.4%) | 3 (8.3%) | 25 (25.0%) |
| Breathing difficulties | 28 (43.8%) | 0 (0%) | 28 (28.0%) |
| Secretion of phlegm | 21 (32.8%) | 2 (5.6%) | 23 (23.0%) |
| **Viruses, n (%)** |  |  |  |
| Rhino/entero | 17 (26.6%) | 2 (5.6%) | 19 (19.0%) |
| Metapneumovirus | 6 (9.4%) | 0 (0%) | 6 (6.0%) |
| Influenza A | 6 (9.4%) | 0 (0%) | 6 (6.0%) |
| Parainfluenza | 3 (4.7%) | 1 (2.8%) | 4 (4.0%) |
| RSV | 3 (4.7%) | 0 (0%) | 3 (3.0%) |
| Adenovirus | 1 (1.6%) | 1 (2.8%) | 2 (2.0%) |
| Bocavirus | 1 (1.6%) | 1 (2.8%) | 2 (2.0%) |
| SARS-CoV-2 | 2 (3.1%) | 0 (0%) | 2 (2.0%) |
| Negative | 7 (10.9%) | 8 (22.2%) | 15 (15.0%) |
| Testing was not done | 21 (32.8%) | 23 (63.9%) | 44 (44.0%) |
| **Hospitalization, n (%)** |  |  |  |
| Hospitalized | 14 (21.9%) | 8 (22.2%) | 22 (22.0%) |
| Not hospitalized | 46 (71.9%) | 28 (77.8%) | 74 (74.0%) |
| Missing<sup>b</sup> | 4 (6.3%) | 0 (0%) | 4 (4.0%) |

N, number; SD, standard deviation; <sup>a</sup>Some patients tested positive for multiple viruses; <sup>b</sup>No permission to access medical records

**Table 2.** Performance of the video-based measurements of respiratory rate, heart rate, and oxygen saturation against clinical standard references in the study population of 64 children with a respiratory infection (cases).

| | Mean (SD) | MAE ± SD | MAPE | RMSE | R<sup>2</sup> |
| --- | --- | --- | --- | --- | --- |
| Respiratory rate (rpm), n=64 | 42.3 (11.3) | 5.4 ± 5.6 | 12.3% | 7.8 | 0.549 |
| Heart rate (bpm), n=48 | 139.3 (24.4) | 16.9 ± 11.0 | 12.6% | 20.1 | 0.339 |
| SpO<sub>2</sub> (%), n=60 | 96.9 (2.3) | 1.2 ± 1.3 | 1.3% | 1.7 | 0.384 |

MAE, Mean Absolute Error; MAPE, Mean Absolute Percentage Error; RMSE, Root Mean Square Error; SD, standard deviation; R<sup>2</sup>, coefficient of determination. Measurement information missing for 16 cases from the HR computation and 4 cases from the SpO<sub>2</sub> computation.

The mean age of the study population was 2.4 years, with a slightly younger mean age observed in the case group (2.2 years) compared to the control group (2.6 years). Boys were overrepresented in the case group (57.8%) compared to the control group (33. 3%), resulting in a general gender distribution of 49% boys and 51% girls throughout this observational case-control study. The children in the case group exhibited significantly higher mean RR and HR compared to the control group. The mean RR was 42.3 breaths per minute (bpm) in cases versus 32.2 bpm in controls, while the mean HR was 139.3 bpm in cases compared to 123.3 bpm in controls. These elevated vital signs are consistent with the clinical presentation of respiratory infections.

Cough, sneezing, breathing difficulties, and phlegm secretion were reported more frequently among cases. Specifically, 56.2% of the cases had a cough compared to just 2.8% of controls, and 43.8% of cases exhibited breathing difficulties, a symptom absent in the control group. Rhinovirus was identified as the most common viral pathogen in the case group (26.6%), followed by metapneumovirus and influenza A (both at 9.4%). Viral testing revealed a higher proportion of negative results among controls (22.2%) than among cases (10.9%). A notable 32.8% of cases did not undergo viral testing, compared to 63.9% of the controls. Hospitalization rates were similar between the two groups, with 21.9% of cases and 22.2% of controls that required admission. However, it is important to note that cases were more likely to exhibit clinical features consistent with respiratory distress, as reflected by higher mean scores for RDAI and PEWS. The mean RDAI score was 2.47 in cases versus 0.06 in controls, while the mean PEWS score was 1.72 in cases compared to 0.29 in controls. The prevalence of asthma was low in the study population, with only one confirmed case (1.6%) among the cases and none in the control group. A small percentage of children had asthma-like symptoms not confirmed with spirometry (4.7% in cases and 5.6% in controls). Maternal smoking was reported in 6.3% of cases and 5.7% of controls, with no significant differences between the groups.

The observed clinical differences between cases and controls, particularly in RR and HR, underscore the importance of accurate measurement and monitoring in children with suspected respiratory infections. The higher prevalence of respiratory distress symptoms and elevated clinical scores among cases highlights the potential to use video-based approaches to identify children with significant respiratory compromise. Variation in viral test rates between groups reflects real-world diagnostic practices but may limit the ability to fully characterize the infectious etiology in some patients.

This detailed analysis provides a comprehensive overview of the study population and serves as a foundation for understanding the performance of video-based diagnostic tools in identifying respiratory infections and related conditions.

### 3.2. Experimental Results

#### 3.2.1. Performance of the computer vision-based vital sign estimates with standard reference in children with respiratory infection (cases)

We estimated three vital signs, including RR, HR, and SpO2, using machine learning in children with respiratory infections (cases). We estimated the three vital signs by training regression models directly on the feature vectors extracted from the video recordings. Each model received as input the features computed from the segmented signal windows. We excluded 16 cases from the HR computation and 4 cases from the SpO2 computation, due to the lack of measurements.

Table 2 presents the results of the video-based estimation of respiratory rate, heart rate, and oxygen saturation compared to the clinical standard. The respiratory rate estimation achieved the highest agreement, with a coefficient of determination of 0.549 and a MAE of 5.4 breaths per minute, corresponding to a MAPE of 12.3%. Heart rate estimation showed a larger variability, with an MAE of 16.9 beats per minute and an MAPE of 12.6%, and a lower determination coefficient of 0.339. This result is consistent with the known sensitivity of rPPG signals to motion and noise, especially in younger patients. Estimation of oxygen saturation produced a small absolute error, with an MAE of 1.2 percentage points and an MAPE of 1.3%, although the explained variance was limited, with a coefficient of determination of 0.384. This could be partially attributed to the narrow distribution of oxygen saturation values observed in the study population. The results show that respiratory rate estimation provided more consistent predictions compared to heart rate and oxygen saturation, reflecting both the potential and limitations of video-based vital sign estimation in pediatric emergency care.

In addition to the general analysis, we performed the analyzes separately for the cases and controls and among the cases stratified by age (0-2 years, 2-5 years) and sex. We report the MAPE results for all data in Table 3.

**Table 3.** Performance of the video-based measurements of respiratory rate, heart rate, and oxygen saturation against clinical standard references in cases, subgroups of cases, controls, and all. Stratified Mean Absolute Percentage Error (MAPE) analysis is used as the performance measure.

| Group | RR (%) | HR (%) | SpO<sub>2</sub> (%) |
| --- | --- | --- | --- |
| Cases (n=64) | 12.3 | 12.6 | 1.3 |
| Age [0 to <2 years) | 12.2 | 13.0 | 1.0 |
| Age [2 to 5 years] | 12.5 | 12.4 | 1.5 |
| Girls | 12.3 | 11.5 | 1.1 |
| Boys | 12.3 | 13.5 | 1.4 |
| Controls (n=36) | 14.9 | 18.4 | 1.0 |
| Total | 13.2 | 14.0 | 1.2 |

The stratified results presented in Table 3 show that, in general, the model performed better in the case group compared to the controls across all vital signs. In particular, both the respiratory rate and heart rate estimations exhibited lower MAPE values in the case group (12.3% and 12.6%, respectively) than in the control group (14.9% and 18.4%, respectively). This difference may be related to the fact that children with respiratory infections tend to remain more still, allowing for more stable signal extraction. Within the case group, the performance was relatively stable across age and sex subgroups, with slightly lower MAPE for heart rate estimation in girls (11.5%) compared to boys (13.5%). The estimation of oxygen saturation consistently yielded low MAPE values in all subgroups, probably due to the narrower variability of this parameter in the study population.

#### 3.2.2. Estimation of pediatric scores

Using a similar approach, we classified the case subjects based on their predicted PEWS and RDAI, as depicted in Figure 4. We divided subjects into three different PEWS score groups: Group 0 (scores between 0 to 3), Group 1 (scores from 4 to 6), and Group 2 (scores of 7 and above). Similarly, subjects were divided into three RDAI score groups: Group 0 (scores between 0 to 4, indicating mild distress), Group 1 (scores from 5 to 8, indicating moderate distress) and Group 2 (scores from 9 to 17, indicating severe distress). Figure 4 shows the confusion matrices for both the classification of the PEWS and RDAI groups.

**Figure 4.**
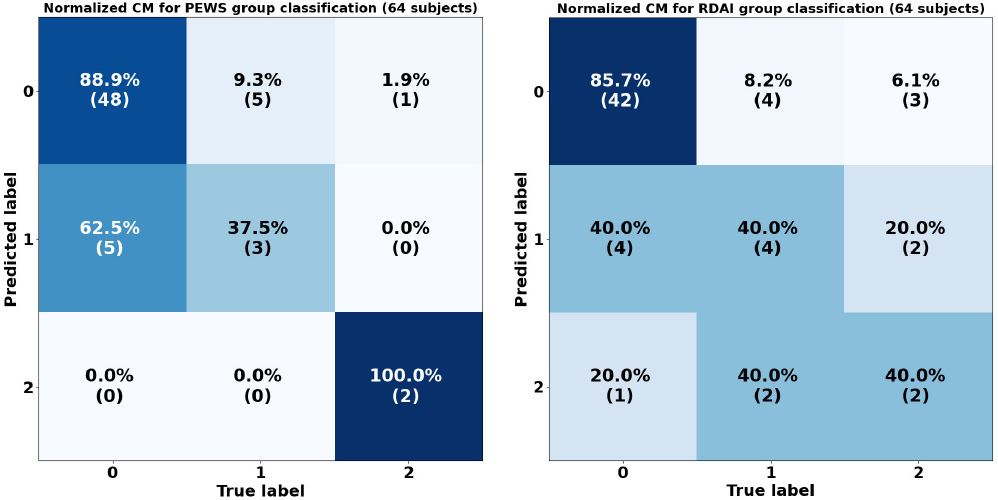
Confusion matrices for PEWS and RDAI estimation for cases.

The analysis of the confusion matrices suggests that the classifiers are highly accurate in classifying subjects in Group 0 with 88.9% and 85.7% accuracies for PEWS and RDAI, respectively. For Group 1, the classifiers only achieved moderate accuracies of 37.5% and 40%, with most misclassifications incorrectly placed in Group 0. The classification of the most severe cases, belonging to Group 2, surprisingly, despite the limited sample size, our classifier achieved a 100% accuracy rate for Group 2 in PEWS, the highest risk group. However, the total accuracy and balanced accuracy stood at 82.9% and 75.5% for PEWS and respectively 79.5% and 55.2% for RDAI.

#### 3.2.3. Prediction of SpO2 group, respiratory infection and hospitalization

We also performed binary classification models developed for the estimation of respiratory infection, the prediction of the need for hospitalization, and the classification of the subjects based on their SpO2. The performance of the model is evaluated using balanced accuracy, a suitable metric for binary classification problems with heavily imbalanced datasets [25], as in these particular tasks.

As an important factor in predicting diagnosis and hospitalization, we performed a binary classification of cases based on two groups, those with SpO2 levels below 95% (range of [90, 95)) and those with SpO2 levels at or above 95% (range of [95, 100]). In this heavily unbalanced scenario, the classifier predicted with an accuracy of 85.7% for subjects having SpO2 levels above 95 and a 72.7% accuracy for those below 95, as depicted in Figure 5a.

**Figure 5.**
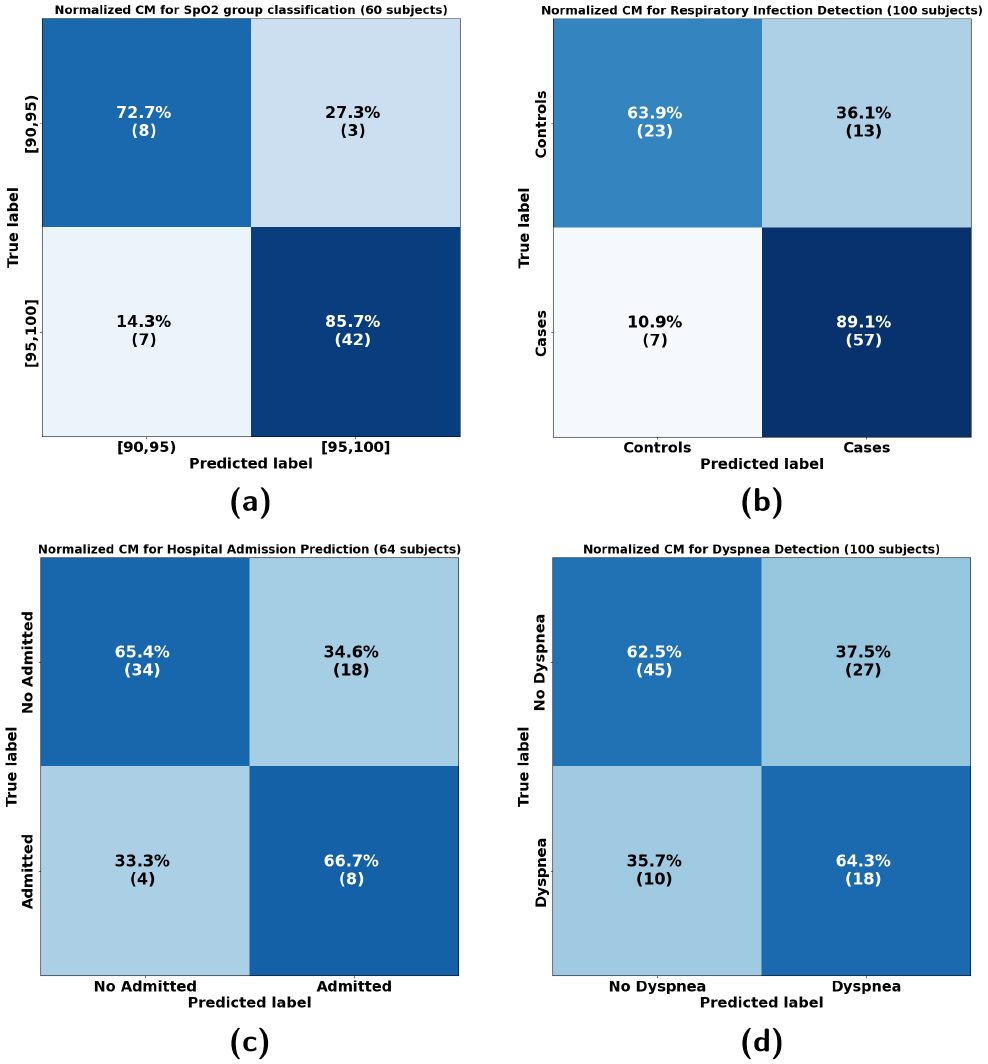
confusion matrices for various classification tasks within the database. Subfigure (a) shows the confusion matrix for SpO2 group classification in the CASES subset. Subfigure (b) illustrates the confusion matrix for Respiratory Infection classification across all subjects in the database. Subfigure (c) depicts the confusion matrix for hospital admission prediction within the CASES subset. Lastly, subfigure (d) displays the confusion matrix for Dyspnea classification across all subjects in the database.

Following on this, for the prediction of diagnosis, we experimented with various thresholds for classifying a patient as case, based on the proportion of windows that the model identifies as indicative of an infection. The best balanced accuracy (76.5%) was obtained at a threshold of 50%, meaning a patient is classified as case if at least 50% of the windows are predicted as infected, but the results only varied slightly across different thresholds. Figure 5b presents the confusion matrix, depicting the performance of the respiratory infection detection model.

Analogously, to predict whether a patient required hospital admission, we obtained a 66.7% of accuracy when at least 35% of the windows are predicted as such, as depicted in Figure 5c. Total balanced accuracy of 66.0%.

#### 3.2.4. Dyspnea Binary Classification in cases

In our study, dyspnea is identified as a critical symptom for binary classification among cases. Similarly to the methods used for SpO2 group prediction and respiratory infection detection, we applied binary classification models to differentiate patients based on the presence or absence of dyspnea. The classification performance is evaluated using balanced accuracy, considering the imbalanced nature of the dataset in this category.

Patients were divided into two groups based on the presence or absence of dyspnea. A predictive model was used to identify patients with dyspnea. The model achieved an accuracy of 64.3% in identifying patients with dyspnea, using a threshold where at least 40% of observed video must indicate dyspnea. For patients without dyspnea, the accuracy was 62.5%. The overall balanced accuracy of the model, considering both groups, was 63.39%. These results are illustrated in Figure 5d.

## 4. Discussion

### 4.1. Main Findings

Our findings indicate a moderate accuracy in estimation all three vital signs via video recordings, suggesting potential applicability in specific clinical contexts. We found an 85.7% accuracy in predicting SpO2 levels above 95 and a 72.7% accuracy for those below 95. The model achieved a 64.3% accuracy in identifying patients with dyspnea and a 62.5% accuracy for those without. The overall balanced accuracy, accounting for both groups, was 63.4%. Regarding pediatric scores, the total and balanced accuracies were 82.9% and 75.5% for PEWS, and 79.5% and 55.2% for RDAI. We obtained a 76.5% accuracy in predicting a patient’s respiratory infection, and a 66.7% accuracy in predicting whether a patient required hospitalization.

The study also suggests that there are only modest differences in accuracy between different age groups and genders, which suggests that single models could be used under relatively general conditions. Notably, both the RR and HR errors are higher for the control group, which could be due to the different causes that brought them to the emergency room. Elevated heart rate error rates in the age category of 2-5 years could be associated with infant tracking complexities, due to unpredictable movements. The slight difference in error rates between girls and boys may suggest small gender-specific variations, supporting previous research that indicates differential activity patterns [27].

Our results indicate that, while rPPG may not provide the necessary resolution for precise SpO2 estimation, it can be leveraged effectively to identify potentially dangerous SpO2 levels when used within an appropriate framework. This finding underscores the potential utility of rPPG in scenarios where traditional pulse oximetry may not be feasible, providing an additional layer of information about the individual’s physiological state.

This application could have the potential to be used in remote medicine, as well as in primary care, hospitals, and other health-care settings. Therefore, it is necessary to investigate its effectiveness in calculating the respiratory rate and obstructive breathing. If computer vision can be used as a tool in healthcare in the future, especially with simple technical devices such as smartphones, the patients in need of urgent care could potentially be separated from healthier patients before even arriving at the emergency department.

### 4.2. Validity of results

#### 4.2.1. Selection bias

The response rate for this study was relatively good (74%). Reasons for declining participation included, for example, the child being too sick, or the parents being tired, the study prolonging the ER visit, and parents’ concern about the nature of the study (the child’s face showing in the video). Therefore, the severity of illness could have affected the parents’ decision whether to participate or not. In addition, selection bias could occur due to the patients being only native Finnish speakers. Due to the language barrier, the study group consisted mostly of Caucasian children with similar skin tone. This should be considered in potential future studies; the application and AI needs to be tested on larger population with more variety.

Age did not affect the selection bias significantly because we approached all children, and then found out if the child was suitable to participate age-wise. Social or economic background could have affected in the less severe cases of illness because we did not include patients from private clinics. In severe cases this does not affect the process of selection due to Finland’s health care system, as all critically ill patients are treated in public health care (University hospital).

#### 4.2.2. Considerable factors in video recording

The objective was to obtain two videos of each patient, which was achieved for 83 participants. There were 23 patients with only one video recorded, 14 sitting and 9 lying down. One patient did not have any video data. Smartphone is a convenient device to be used in the emergency department, and most children in Finland are used to smartphones as opposed to a bigger video camera. The problems that occurred in the video data gathering process were related to the young age of the patients as they were not able or patient enough to sit by the protocol and therefore only a video laying down was able to be taken. On the other hand, the older age group was more difficult to motivate to the study and to stay still, which might be why we extracted better signals from younger children. Some children were also afraid and crying of the video-taking or of the clinical examination situation, started focusing on their breathing too much or holding their breath, which could lead to difficulties in the video data analysing process. Some children had a pacifier, clothes they refused to take off or a toy in the way, which also could make video analysis problematic. Boys were also moving more that girls according to our findings.

The mobility of the child was an important factor in the successful recording of the video. In most cases, the children of the case group were sicker than the controls, as measured with previously mentioned PEWS- and RDAI-scores. This could have affected the results, as the sicker children remained still and had more visible breathing. Therefore, their signals might have been easier for the computer vision to interpret.

In this study, all videos were recorded in similar lighting and with the same camera. In the future, the application and AI needs to be tested in different situations to ensure the function in different environments or make the baseline on what the conditions for video recording need to be.

#### 4.2.3. Evaluation by Doctors

The doctors in the ER change almost daily and therefore many different doctors filled out the health data form, and subjectively assessed the patient’s state, which could lead to differences in perspective when filling out the health form (especially the RDAI score). The RR was measured by different professionals and under different conditions. In most cases, the RR was measured by a nurse prior to the clinical examination of the doctor, and the time between measurements and the evaluation of the doctor and the video recording varied from minutes to hours. In some cases, the RR was measured by nurses prior to giving the patient salbutamol [28], and the video was taken afterwards. In these cases, we re-evaluated the RR. Sometimes, the RR was an estimate made by doctors and not accurately measured. In these cases, we also re-evaluated the RR.

With some patients, the health form did not have all the measurements, for example HR or SpO2, which was mostly the case with some control patients. This could lead to difficulties in assessing the PEWS-score. The RDAI-score was complete with all the patients.

### 4.3. Synthesis with previous knowledge

In the field of non-contact vital sign monitoring, previous studies have varied greatly, particularly when considering different subject age groups and their respective environments. Most studies have focused on adult populations, where they have typically focused on static settings, such as sleep monitoring. The accuracies reported for these settings are close to those of other medically graded measurement devices, when the measurement conditions are stable and controlled. For example, Zhu et al. (2019) [29] demonstrated accuracy in respiratory rate estimation with around 2 breaths per minute of error during most of the monitoring time (about 90% of the time). Villarroel et al. (2019) reported a MAE of 2.8 bpm for heart rate and 2.1 breaths/min for respiratory time, but valid only during less of 76% of the measuring time [30]. Bae et al. (2022) measured vital signs from facial videos collected from healthy collaborative adults, and achieved a respiratory rate MAE below 1 breaths/min [31].

In contrast, studies focusing on neonatal and infant populations have more varied results, although they have mostly been performed in controlled environments. For newborns in the NICU, Villarroel et al. (2014) achieved a heart rate MAE of 2.8 bpm and claimed that comparable results could be expected for respiratory rate [32]. However, their system was effective in the measurement during 52% of the time. Lorato et al. (2021) showed an MAE of 3.3 breaths/min on the testing set and 5.4 breaths/min on the validation set, while their system was applicable for 64.0% and 69.7% of video segments, respectively [33]. Rossol et al. (2020) reported a significant correlation between camera-based measurements and standard ECG impedance pneumography, with an RMSE error exceeding 6 breaths per minute [34]. In a different setting, Cañada Carril et al. (2023) tested a camera setting for the detection of respiratory rate in children under 10 years of age with respiratory distress. The patient’s chest movement was controlled with two green stickers on the body, easily trackable for the camera. In their preprint, they reported an MAE between 6.7 and 8.9 breaths / min [35].

Our study differs fundamentally from previous approaches. Located in a pediatric emergency department, our environment is far more dynamic and varied than the controlled settings of other studies. We focused not only on measuring vital signs but also on identifying respiratory infection or distress in children. Using machine vision to estimate vital signs (RR, HR, SpO2), our approach caters to a broader range of vital signs than most studies, which typically focus on one or two parameters. Additionally, our study acknowledges the high standard deviation and variability across subjects, highlighting the challenges in achieving consistent accuracy in a real-world, high-variability clinical environment.

### 4.4. Clinical significance

Our study found relatively high accuracy for predicting a patients’ respiratory infections. Our results demonstrated that all three vital signs (HR, SpO2 and RR) can be estimated with moderate accuracy, which could be considered acceptable in various scenarios. However, a relatively high standard deviation across subjects suggests that the reliability of the measurements could still be improved. Alternatively, identifying the technical, medical, and environmental factors contributing to higher errors should be explored more thoroughly. An important aspect to consider is that the ground truth data was not recorded simultaneously with the video recordings, adding an additional layer of complexity to the task. Vital signs subject to fast variation, such as HR and RR, are likely to vary slightly in a short period of time. Consequently, this should be considered when interpreting the results.

When comparing the predictive outcomes between the RDAI and PEWS scores, results indicated a marginally superior performance with PEWS. This disparity could arise due to the RDAI score being heavily reliant on meticulous evaluation of the patient’s respiratory patterns, beyond the respiratory rate. Given the considerable disparities present in our dataset, it is imperative to recognize that while these findings are promising, they need additional in-depth analysis and verification to reach definitive conclusions.

Although the results are moderately good, the imbalance of the dataset suggests that more data is needed to accurately predict the admissions and infections, since misclassifications in these are more relevant.

## 5. Conclusion

This study demonstrates the potential of computer vision systems to complement traditional diagnostic workflows in pediatric emergency care by providing automated, non-intrusive assessments of respiratory conditions. While the results show moderate accuracy, further research is needed to improve reliability across diverse clinical scenarios and patient populations.

The primary objective of this study was to evaluate the effectiveness of using a novel computer vision and signal processing approach for extracting physiological biosignals via a mobile phone camera in an unsupervised setting. This innovative method uses rPPG to capture blood volume pulse from subtle skin color changes, and rBSG to detect bodily movements that indicate respiratory activity and pediatric patient restlessness. By integrating these techniques, we could reliably compute vital signs such as HR, RR, and SpO2, which are crucial for assessing the respiratory health of patients.

Additionally, our study extended to analyze HRV and respiratory rate variability (RRV), which are significant markers of respiratory distress. These metrics contributed to the calculation of pediatric medical scores, like the PEWS, enhancing the clinical assessment capabilities for pediatric patients. The analysis also utilized these indicators to train models for classifying potential infections, thereby providing a basis for diagnostic support that is not only effective but also interpretable to healthcare professionals.

While our findings demonstrate the significant potential of contactless diagnostic tools in pediatric care, the results also highlight the necessity for broader data collection and further experimentation. The limited sample size of 100 patients introduces a degree of bias that could impact the reliability of the outcomes. It is imperative that future research efforts focus on expanding the dataset and refining this methodology to ensure robust validation and enhance the accuracy of these promising diagnostic approaches. This will be critical for facilitating their integration into clinical practice, where they can offer rapid, non-invasive assessments, crucial for effective pediatric care.

## Data Availability

Due to the sensitive nature of the data, including video recordings of pediatric patients, and the restrictions imposed by the ethical approval obtained from the Regional Medical Research Ethics Committee of the Wellbeing Services County of North Ostrobothnia, Finland, the data cannot be shared publicly. The pseudonymized data are stored on encrypted servers within the secure infrastructure of Oulu University Hospital and are not available for external use.

## Ethical Statement

The study protocol was approved by the Regional Medical Research Ethics Committee of the Wellbeing Services County of North Ostrobothnia, Finland. This was a population-based incident case-control study conducted at the Emergency Department of Oulu University Hospital, which serves all acute pediatric cases from a geographically defined region in Northern Finland. The recruitment took place between April and June 2022.

Children aged 0–5 years and their guardians were invited to participate upon arrival at the Emergency Department. Non-Finnish speakers were excluded. Written informed consent was obtained from all parents or legal guardians prior to participation. The study adhered to relevant ethical regulations and guidelines for research involving human participants. All collected data were handled confidentially and anonymized prior to analysis.

## Acknowledgment

The research was supported by 6G Flagship (Grant Number 369116) funded by the Research Council of Finland, PROFI5 Hi-Dyn (Grant Number: 326291), and the Orion Research Foundation (Grant: Apurahat Constantino Álvarez Casado). We also acknowledge Guillermo Jimenez and Eduardo Gracidas, authors of the Rho LaTeX template (Version 2.1.1), which is distributed under the Creative Commons CC BY 4.0 license, for providing the template used to prepare this manuscript. Illustrative elements in Figure 2 were generated using OpenAI’s ChatGPT-4o for schematic visualization purposes.

## Supplementary Material

This section provides additional figures, tables, or explanations that support the main text.

## Methodology for Pediatric Support Medical Diagnosis via Computer Vision, Signal Analysis, and Machine Learning

Our approach to support pediatric medical care involves a comprehensive automatic analysis of recorded videos. We leverage state-of-the-art computer vision and signal processing techniques to extract respiratory and heart biosignals from the face and torso of patients, and machine learning to classify respiratory conditions using computed respiratory and heart-related features extracted from the biosignals.

The video analysis methodology consists of five main stages, as depicted in Figure 2. First, we capture high-quality video streams of patients using the rear camera of a smartphone. Second, we perform face and body detection, tracking, and skin segmentation to isolate the relevant regions of the face and precordium. For facial analysis, we follow a similar approach as Alvarez Casado et al. [24]. We used a deep learning-based face detection method based on the Single Shot Multibox Detector (SSD) network [36], implemented with the *OpenCV* library. Compared to traditional detectors, this approach offers a good balance between accuracy, computational speed, and model size [37]. For face alignment, we applied the Deep Alignment Network (DAN) [38], which predicts 68 facial landmarks following the Multi-PIE convention and shows robust performance even under challenging conditions [39]. The segmented facial region was further refined using a geometric skin segmentation approach based on landmark positions, as proposed by Álvarez Casado et al. [24]. For body analysis, we used the Detectron2 library [40] by Facebook AI Research to detect the precordium region, perform keypoint estimation, and track body movements. Detectron2 enables robust segmentation and localization of the upper torso, allowing us to extract motion-based biosignals and also allowing for skin segmentation. This combination of face and body analysis provided the basis for computing both rPPG and rBSG signals used in the vital sign estimation process. Third, we extracted a total of four biosignals from these regions by analyzing subtle variations in color (remote photoplethysmography) or motion (remote ballistography). By analyzing both color variations and movements in the face and precordium regions, we obtain three groups of raw signals: *raw_rgb_face, raw_rgb_thorax, raw_mov_face, and raw_mov_thorax*. To extract the physiological signals, we first processed the color and motion signals obtained from the video. The raw RGB color signals were converted into rPPG signals using the OMIT matrix decomposition method [24], which enhances the extraction of pulse-related variations. For motion signals, we computed the total body movement by calculating the Eulerian distance (L2 norm) between consecutive keypoints, resulting in rBCG signals. Both signals were then filtered using bandpass filters: one tuned to the heart rate frequency band (0.75–4 Hz, corresponding to 45–240 beats per minute) and another tuned to the respiratory rate band (0.1–1.4 Hz, corresponding to 6–84 respirations per minute). This filtering step isolated the relevant cardiac and respiratory components, which were then used for feature extraction, as illustrated in Figure 2.

The next step involves the computation of vital signs and the extraction of features from the processed biosignals. The extracted signals, including rPPG and rBCG, are segmented into fixed-length windows for analysis. Specifically, each signal is divided into windows of 11 seconds with a sliding step of 1 second, resulting in approximately 18 to 25 windows per video, depending on its length. This overlap (90%) ensures temporal continuity and robustness in feature extraction. For each window, we computed a total of 250 features derived from the three one-dimensional biosignals. The extracted features include statistical descriptors, heart rate variability (HRV) and respiratory pattern indicators, morphological characteristics, and descriptors of complexity, entropy, and fractal properties. Statistical features consist of common signal descriptors, such as mean, minimum, maximum, standard deviation, dynamic range, signal-to-noise ratio (SNR), median, and percentiles (10th, 25th, 75th, and 90th). Fractal analysis features include the Katz fractal dimension, Higuchi fractal dimension, and detrended fluctuation analysis, computed for the entire window, and averaged over four non-overlapping sub-windows of 3 seconds. Entropy-based features consist of permutation entropy, spectral entropy, approximate entropy, sample entropy, Hjorth mobility and complexity, and the number of zero-crossings. Complexity-related measures include complexity tolerance, complexity dimension, and complexity delay. For heart and HRV-related features, we computed up to 40 features such as heart rate (HR), breathing rate (BR), inter-beat interval (IBI), pNN20 and pNN50 (proportions of successive R-R intervals differing by more than 20 ms and 50 ms, respectively), Poincaré descriptors, frequency-domain features (very low frequency, VLF; low frequency, LF; high frequency, HF; LF/HF ratio), and the standard deviation of normal-to-normal intervals (SDNN). The implementation of these calculations was performed using Python libraries. Statistical features were computed using the Numpy library, entropy and fractal features were obtained with the Antropy package [41], and HRV-related features were extracted using NeuroKit2 [42] and HeartPy [43]. Furthermore, we included handcrafted features to describe respiratory dynamics, such as peak-to-peak mean distance, standard deviation of inter-inhalation intervals, exhalation duration, and the differential between rPPG signals obtained from the face and thorax. These feature vectors were then used as input for classification models to detect respiratory conditions and for regression models to estimate relevant vital signs and physiological parameters.

The extracted features were subsequently used to train several machine learning models, including XGBoost, Support Vector Machine (SVM), Random Forest (RF), and Multi-Layer Perceptron (MLP), using the Scikit-learn and XGBoost Python libraries. These models were applied to both classification and regression tasks. The classification tasks focused on identifying the presence of respiratory infection and determining whether hospitalization was required. Regression tasks involved estimating oxygen saturation (SpO2), Pediatric Early Warning Score (PEWS), Respiratory Distress Assessment Index (RDAI), respiratory rate, and heart rate. Among the tested models, XGBoost consistently yielded the most reliable results across both classification and regression problems, followed by Random Forest. The superior performance of XG-Boost is likely attributed to its ability to handle heterogeneous feature sets, control overfitting through regularization, and its suitability for structured, tabular data with mixed types of features, as is typical in biomedical signal processing. Therefore, XGBoost was selected as the main model for reporting results, while the other models served for comparison and sensitivity analysis.

## Data Collection and Management

After the collection of a patient’s data, which included paperwork and videos, the documents were securely stored in a locked office within the OYS facility. The videos were saved onto an encrypted solid-state drive, also kept in the locked office. The study’s progress was recorded in a logbook, also kept in the secure location, which documented the patient’s identification information, respiratory status, and which videos were taken.

The personal information was stored in an Excel file, and the anonymous data was stored in a separate Excel file. The anonymous data was then transferred to SPSS statistical software for analysis. This strict data management protocol ensured the confidentiality and security of the patient’s information throughout the entire study.

## Video Analysis

The videos were evaluated by our study’s technical team from the Center for Machine Vision and Signal Analysis (CMVS), and they were divided into three categories: perfect, good enough and questionable usability/not usable. Perfect meaning both videos are recorded, and the quality is good, and the information of the patient contains respiratory rate. Good enough meaning at least one video is recorded and quality is good enough, and the information of the patient contains respiratory rate. Questionable usability/not usable meaning neither of the videos are usable (due to back illumination, occlusion caused by clothes or a pacifier, or bad positioning), there are no videos, or the subject information does not contain respiratory rate. There were 51 perfect videos, 37 good enough videos and 13 patients with videos of questionable usability.

